# Next-Generation Sequencing-Based Analysis of HLA Variants in Turkish Patients with Obstructive Sleep Apnea

**DOI:** 10.1101/2021.06.03.21258051

**Authors:** Ayse Gul Zamani, Sebnem Yosunkaya, Celalettin Korkmaz, Adil Zamani, Hulya Vatansev, Ahmet Burak Arslan, Mahmut Selman Yildirim

## Abstract

**Background:** Obstructive sleep apnea (OSA) is a highly prevalent, complex respiratory disorder characterized by repetitive episodes of upper airway collapse. While its pathogenesis involves various anatomical and neuromuscular factors, the underlying genetic architecture remains partially elucidated. The human leukocyte antigen (HLA) system, located on chromosome 6p21, is the most polymorphic region of the human genome and plays a pivotal role in immune regulation and inflammatory pathways, which are often implicated in OSA. Historically, HLA studies in OSA relied on lower-resolution serological methods, often yielding inconsistent results. Recent advancements in targeted Next-Generation Sequencing (NGS) now provide an unprecedented opportunity to analyze HLA alleles at the four-to-six-digit level, offering the precision required to identify specific genetic markers.

**Aims:** The primary objective of this study was to conduct a high-resolution analysis of genetic variants at the HLA-A, -B, -C, -DQB1, and -DRB1 loci using targeted NGS technology to identify predisposing or protective alleles in Turkish patients with OSA compared to a healthy cohort.Study

**Design:** A prospective case-control study.

**Methods:** The study included 100 participants, consisting of 50 newly diagnosed patients with OSA and 50 healthy controls. The diagnosis of OSA was confirmed via comprehensive overnight polysomnography, and patients were categorized based on an apnea-hypopnea index of five or more events per hour. Control subjects were prospectively selected from volunteers with a low clinical risk for OSA and normal sleep patterns. High-resolution genotyping of the major HLA loci was performed using an Illumina-based targeted NGS platform. Statistical analyses were conducted to compare allele frequencies and identify significant associations between specific HLA variants and disease susceptibility.

**Results:** High-resolution NGS analysis revealed significant associations for several HLA alleles. HLA-A*02:01, HLA-C*03:03:01, HLA-C*14:03, and HLA-DRB1*04:05 alleles were found to be significantly more frequent in the OSA group compared to the control group (p=0.036, p=0.007, p=0.043, and p=0.013, respectively), suggesting a potential predisposing role. Conversely, the frequencies of HLA-A*03:01 (p=0.024) and HLA-B*35:02 (p=0.043) were significantly higher in the control cohort, suggesting that these alleles might confer a protective effect against the development of OSA.

**Conclusion:** Our findings suggest that HLA-A*02:01, HLA-C*03:03:01, HLA-C*14:03, and HLA-DRB1*04:05 alleles may serve as predisposing genetic markers for OSA in the Turkish population. The utilization of high-resolution NGS provides a more definitive genetic profile than previous methodologies. These findings contribute to the growing body of evidence regarding the immunogenetic basis of OSA and may facilitate the identification of high-risk individuals through genetic screening in the future.

## INTRODUCTION

Obstructive sleep apnea (OSA) is among the most prevalent of sleep-disordered breathing (SDB) types, characterized by recurrent episodes of partial or complete collapse of the upper airway during sleep, resulting in reduced (hypopnea) or absent (apnea) airflow lasting for at least 10 seconds and associated with either cortical arousal or a fall in blood oxygen saturation. The presence and severity of the disease is determined by the amount of respiratory events per hour during sleep: apnea-hypopnea index (AHI).(1) Current epidemiological data indicate that moderate-to-severe OSA affects approximately 17% of middle-aged men and 9% of middle-aged women.(2) While obesity remains the primary modifiable risk factor present in roughly 70% of cases(3) non modifiable factors such as age, male sex, and craniofacial morphology play a significant role in disease development.(4) These are thought to be affected by genetic factors. There is a growing evidence that genetic factors, and their interaction with environmental exposures, influence the development of OSA.(5, 6) Evidence from family-based and twin studies, as well as the association of specific hereditary craniofacial disorders and the Prader-Willi syndrome with OSA, indicate that the disease may have a genetic basis.(5) However, the genetic basis of the disease is yet to be clearly identified.

It has been estimated that approximately 40% of the variance in the AHI may be explained by familial factors. It is likely that genetic factors associated with craniofacial structure, body fat distribution and neural control of the upper airway muscles interact to produce the OSA phenotype.(6) Genetic factors have been reported to play a role in OSA development through pathways involved in ventilatory control, craniofacial anatomy, inflammation, and immunity.(7)

The human leukocyte antigen (HLA) system plays a fundamental role in the regulation of immune responses and the maintenance of self-tolerance by discriminating between self and non-self antigens. The HLA complex, a cluster of genes encoding the major histocompatibility complex (MHC), is located on chromosome 6p21.3 and represents the most polymorphic region of the human genome. Genetic variations within this region have been associated with the pathogenesis of more than 100 diverse conditions, ranging from classic autoimmune disorders like type 1 diabetes and rheumatoid arthritis to chronic inflammatory diseases such as psoriasis and asthma.(8, 9) The HLA genes are functionally categorized into two primary groups: Class I molecules (including HLA-A, -B, and -C) and Class II molecules (including HLA-DRA, -DRB1, -DQA1, -DQB1, -DPA1, and -DPB1). These loci are situated within the MHC region on chromosome 6p21.3, harboring over 20,000 closely related alleles. Both Class I and Class II molecules are essential for antigen presentation, delivering short peptide fragments to T lymphocytes. Specifically, Class I molecules present endogenous peptides ranging from naturally occurring cellular proteins to mutated, damaged, or viral proteins to CD8+ T lymphocytes. Conversely, Class II molecules primarily present exogenously derived peptides, such as bacterial proteins, to CD4+ T lymphocytes.(9, 10) Serological methods are inherently limited, as they can identify only a small fraction of the vast HLA allelic diversity. Alleles with distinct DNA sequences may encode proteins with identical serological reactivity, leading to significant ambiguity. Since the 1980s, the advancement of HLA gene sequencing has facilitated a rapid expansion in the repertoire of newly identified alleles. Consequently, HLA typing has evolved beyond serological reactivity to include precise DNA-based molecular typing and amino acid sequence analysis. While many laboratories globally have implemented sequence-specific oligonucleotides (SSO), sequence-specific primers (SSP), and Sanger sequencing, these methodologies possess inherent constraints. Notably, SSO and SSP techniques are restricted to the detection of previously characterized alleles, limiting their utility in identifying novel genetic variations.(10, 11) Sanger sequencing, while more detailed, is a stepwise process that often fails to resolve certain heterozygous single nucleotide variations (SNVs); furthermore, it remains a costly and cumbersome methodology. HLA alleles are reported on a spectrum ranging from low to high resolution, dictated by the specific typing method employed. Serological typing yields low-resolution results, typically reported at the two-digit level (e.g., HLA-A01 or HLA-A02). While SSO and SSP techniques achieve high accuracy at a four-digit resolution, they are fundamentally limited to known alleles. In contrast, Next-Generation Sequencing (NGS) provides high-resolution data where alleles are defined as unique nucleotide sequences using all defining digits (e.g., A01:01:01:01 and A02:07), thereby enabling detection at the four-to-six-digit level. Consequently, NGS technologies significantly enhance HLA typing speed by facilitating the high-throughput analysis of large sample cohorts with superior resolution and reduced cost per sample.(11)

The aim of the present study was to investigate the genetic variants at the HLA-A, -B, -C, -DQB1, and DRB1 loci in Turkish patients with obstructive sleep apnea and healthy individuals using high resolution targeted next-generation sequencing, in light of these technological advancements.

## MATERIALS AND METHODS

### Study Design and Ethical Statement

This prospective case-control study was conducted at the Department of Pulmonary Medicine and the Department of Medical Genetics of Necmettin Erbakan University, Meram Medical Faculty Hospital. The study protocol was approved by the Institutional Ethics Committee of Necmettin Erbakan University, Meram Medical Faculty (Decision No: 2017/827) and was supported by the Scientific Research Projects Coordination Unit of the same university (Project No: 171218006). All procedures were performed in accordance with the ethical standards of the Declaration of Helsinki. Prior to participation, written informed consent was obtained from all individuals included in the study.

### Participant Characteristics and Control Group Selection

A total of 100 participants were prospectively enrolled, comprising 50 newly diagnosed patients with OSA and 50 healthy control subjects. The control group consisted of 50 unrelated, healthy volunteers who had applied to the tissue typing laboratory as potential transplant donors. These individuals were randomly selected from a cohort undergoing routine clinical and laboratory screening. To ensure a robust control population, subjects were included only if they had no prior medical history of chronic disease or clinical symptoms suggestive of OSA. Furthermore, all participants in the control group completed the Epworth Sleepiness Scale (ESS) questionnaire to objectively exclude excessive daytime sleepiness.(12) Written informed consent was obtained from all participants prior to their inclusion in the study.

### Exclusion Criteria

Patients diagnosed with other sleep disorders, including central sleep apnea, sleep-related hypoxemia, sleep-related hypoventilation, narcolepsy, insomnia, circadian rhythm sleep-wake disorders, and periodic limb movement disorder, were excluded. Additionally, individuals with comorbidities known to be associated with OSA such as Cushing’s syndrome, acromegaly, neuromuscular disorders, and untreated hypothyroidism were not enrolled. To eliminate potential respiratory interference, patients with known severe pulmonary diseases (asthma, COPD, bronchiectasis, etc.), those exhibiting airway obstruction (FEV1/FVC < 70%) on pulmonary function tests according to GOLD criteria, or individuals with any radiographic pathology on chest X-ray were also excluded from the study.(13)13

### Polysomnography Protocol

All patients included in the study underwent comprehensive overnight polysomnography (PSG) using a SOMNOscreen plus (SomnoMedics GmbH, Randersacker, Germany) device in our sleep laboratory. The PSG recordings consisted of a four-channel electroencephalogram (EEG: C3/A2, C4/A1, O1/A2, O2/A1), a two-channel electrooculogram (EOG: right and left), and submental electromyography (EMG). Respiratory monitoring included a nasal cannula and thermistor for airflow, tension-sensitive belts placed on the thorax and abdomen to detect respiratory effort, and a pulse oximeter for oxygen saturation. Additionally, leg EMG (tibialis anterior muscle), a snoring sensor, and position sensors were utilized. The analysis and interpretation of PSG data were performed by a certified sleep specialist in accordance with the American Academy of Sleep Medicine (AASM) standards. Apnea was defined as a reduction in airflow of ninety percent or more from the baseline lasting at least ten seconds. Hypopnea was defined as a reduction in airflow of thirty percent or more accompanied by oxygen desaturation of three percent or more or an arousal for a duration of at least ten seconds.(14)

### Disease Severity and Respiratory Parameters

The apnea-hypopnea index (AHI) was calculated by dividing the total number of apnea and hypopnea episodes by the total sleep time in hours. Disease severity was classified into four categories based on the AHI values: “simple snoring” (AHI 0-5), mild OSA (AHI 5-15), moderate OSA (AHI 16-30), and severe OSA (AHI > 30). In addition to the AHI, the following respiratory and sleep parameters were recorded for each patient:

Minimum oxygen saturation (min SaO2): The lowest oxygen saturation value recorded during sleep. Average oxygen saturation (avSaO2): The mean oxygen saturation level throughout the night.

Time SaO2 < 90%: The percentage of sleep time spent with an oxygen saturation level below 90%. Sleep efficiency (SE): The ratio of total sleep time to total time spent in bed.

REM duration rate (% REM) and NREM3 duration rate (% NREM3): The respective ratios of REM and deep sleep (NREM3) durations to the total sleep time.(15)

All participants, including both patients and controls, were requested to complete the Epworth Sleepiness Scale (ESS) questionnaire during their initial clinical interview. The ESS is a validated instrument used to subjectively evaluate daytime sleepiness by scoring the likelihood of falling asleep in eight distinct daily situations. The Turkish version of the scale, which has been previously validated, was utilized in this study. Participants who obtained a total score of ten or higher were considered to exhibit excessive daytime sleepiness.(16)

### HLA Genotyping by Next-Generation Sequencing

Peripheral blood genomic DNA was extracted using a QIAamp DNA Blood Mini Kit (QIAGEN, Germantown, MD, USA), in strict accordance with the manufacturer’s instructions. The quality and concentration of the extracted DNA were assessed and normalized prior to sequencing. High-resolution HLA typing for the HLA-A, -B, -C, -DRB1, and -DQB1 loci was performed using the MIA FORA NGS FLEX HLA Typing Kit (Immucor Inc., Norcross, GA, USA; formerly Bio Array Solutions Ltd.) on the Illumina MiniSeq (Illumina, San Diego, CA, USA) platform, as previously described.(17)

### Sequencing Coverage and Depth

The genomic coverage for the HLA-A, -B, and -C loci encompassed all exons and introns, including at least 200 base pairs of the 5′ untranslated region (UTR) and 100 to 1100 base pairs of the 3′ UTR. For the -DRB1 locus, coverage included all exons, introns 2 through 6, at least 440 base pairs of the 5′ UTR, 12 base pairs of the 3′ UTR, and specific segments of intron 1 (275 base pairs adjacent to exon 1 and 210 base pairs adjacent to exon 2). The -DQB1 locus coverage included exons 1 through 5 and introns 1 through 4, while the -DPB1 locus included exons 2 through 4 and introns 2 through 3 (Figure 1).(18)

**Figure 1.**
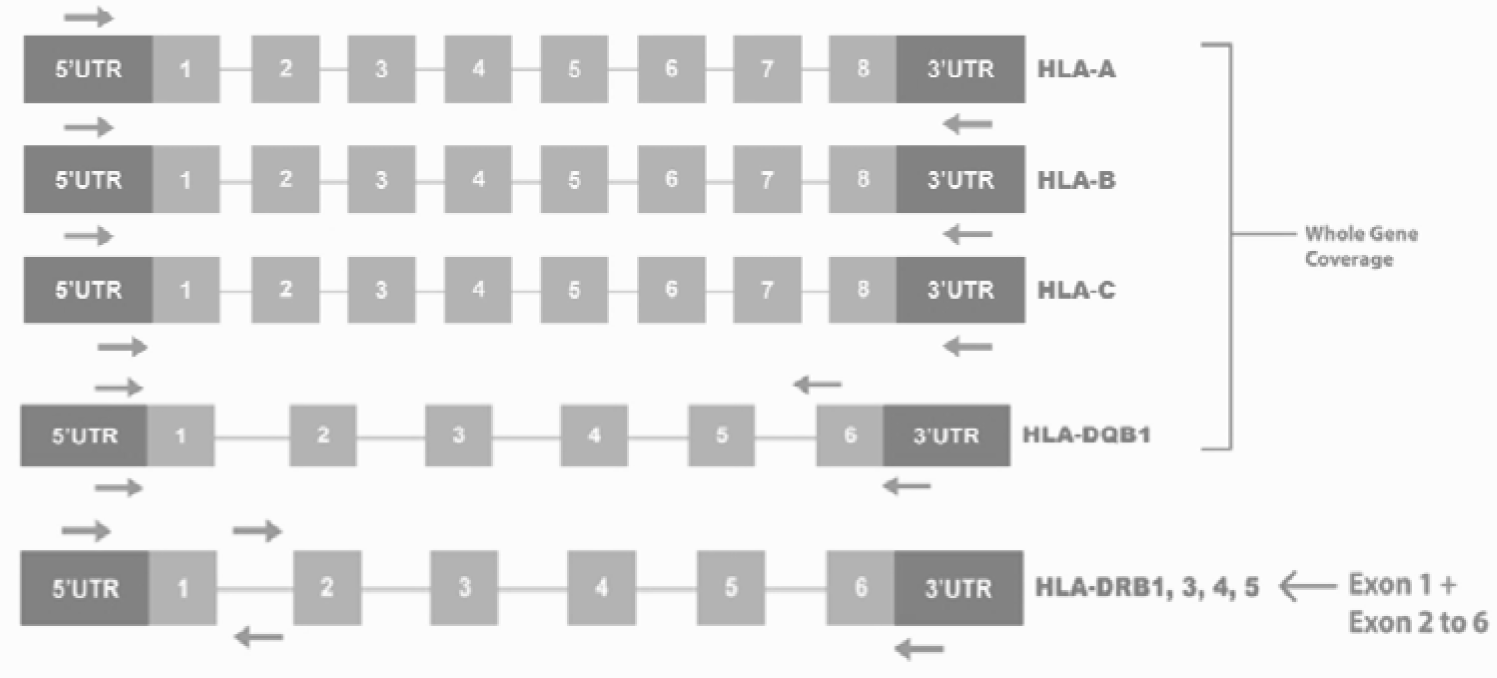
Coverage of HLA loci using MIA FORA NGS HLA typing kit. Whole gene is sequenced for HLA-A, -B, -C and DQB1. DRB1 is sequenced for exon1 and exon2 to exon 6. Courtesy of. https://www.immucor.com/en-us/Products/Pages/ **MIA-FORA-NGS.aspx. UTR, untranslated region.^17^**

### Library Preparation and NGS Data Analysis

In brief, long-range PCR was performed for HLA genes using 5–15 ng/µl of genomic DNA. The concentration of each amplicon was determined using the PicoGreen® assay. Balanced, pooled amplicons were fragmented, end-repaired, and A-tailed. Subsequently, samples were ligated with unique index-adaptors. The pooled, clean, and index-adaptor-ligated samples were purified using the Pippin Prep system (Sage Science, Beverly, MA, USA), which selected DNA fragments within the desired range (500–900 bp). The amplified sequencing library was then cleaned, quantified, and prepared for Illumina sequencing. The average sequencing coverage generated was ≥40×. The resulting sequencing data were analyzed using the MIA FORA NGS FLEX HLA Genotyping Software (version 3.0) with reference to the IPD-IMGT/HLA database (release 3.27.0).(19)

### Bioinformatics and Allele Identification

A mismatch filter was applied to eliminate alignments containing discrepancies or gaps, while a paired-end filter was utilized to enhance specificity by requiring both ends to map to a single HLA reference sequence. Subsequently, the minimum coverage was computed for each candidate allele, and allele pairs were determined according to the methodology described by Wang et al.17 Alleles were identified based on variations within the coding sequences only, ensuring a high-resolution focus on functional genetic diversity.

### Statistical Analysis

Continuous variables were expressed as the mean ± standard deviation. Normality of the data was assessed, and comparisons between groups were performed using the Student’s t-test or chi-square (χ^2^) tests, depending on the distribution and characteristics of the data. All statistical analyses were conducted using the SPSS software (version 22.0; IBM Corp., Armonk, NY, USA).The allele frequencies for the HLA loci were calculated for both the patient and control groups. These frequencies were compared using the χ^2^ test or Fisher’s exact test where appropriate. To determine the strength of the association, Odds Ratios (OR) and 95% Confidence Intervals (CI) were calculated. A p-value of less than 0.05 was considered to indicate statistical significance (p < 0.05).

## RESULTS

The demographic characteristics and ESS scores of OSA patients and normal controls are detailed in Table 1. There was no significant difference between cases and controls group in terms of age, gender and BMI (all p < 0.05). There was only a significant difference between the two groups in terms of ESS scores (p< 0.0001). Polysomnographic data of OSA patients were given in Table 2.

**Table 1.** Demographic characteristics and clinical data of patients with OSA and normal controls.

|  | OSA(n=50) | NC (n=50) | *P* value |
| --- | --- | --- | --- |
| Age(year) | 55.8 ±4.5 | 55.72± 8.9 | 0.95 |
| Male, n (%) | 31(%62) | 31(%62) | NS |
| BMI (kg/m2) | 33.08 ±3.2 | 32.4 ± 1.9 | 0.1994 |
| ESS score | 11.5 ± 4.8 | 7.4 ± 2.4 | < 0.0001 |

Data are expressed as n (%) or mean ± standard deviation. Abbreviations: OSA-obstructive sleep apnea; NC-normal control; NS- non-significant; BMI: body mass index; ESS-Epworth sleepiness scale.

**Table 2.** Polysomnographic data of OSA patients.

| Parameters | mean±SD |
| --- | --- |
| Sleep Efficiency (%) | 81.2 ± 55.1 |
| NREM3(%) | 6.90±1.88 |
| REM(%) | 10.8 ± 11.9 |
| AHI(event/hour) | 28.9 ± 25.6 |
| Minimum SaO<sub>2</sub> | 79.8 ± 10.9 |
| Mean SaO<sub>2</sub> | 89.6 ± 4.4 |
| t<%90 SaO<sub>2</sub> | 34.81±8.54 |

Abbreviations: AHI, apnea-hypopnea index; NREM3, the rate of non-REM sleep time to the total sleep time; REM, the rate of REM sleep time to the total sleep time; SE, the ratio of time spent asleep to time spent in bed; t SaO<sub>2</sub> ≤ %90, the ratio of saturation spent below 90% of sleep time to the total sleep time. Data are expressed as mean ± standard deviation.

Comparative results of patient-control groups for this region are summarized in Table 3. There was a statistically significant difference between these groups in terms of HLA-A * 02: 01 and HLA-A * 03: 01 alleles (p = 0.036, p = 0.024, respectively). HLA-A * 02: 01 was observed more frequently in the patient group, while HLA-A * 03: 01 was more frequent in the control group. It was shown that the HLA-A * 02: 01 allele was susceptible to the disease and the HLA-A * 03: 01 allele was protective. The p value of the HLA-B * 35: 02 allele (p = 0.043)was statistically significant, HLA-B * 44: 03 allele was considered insignificant (p = 0.054). Nevertheless, it should be noted that this is a borderline value. The HLA-B * 35: 02 allele was evaluated as protective. HLA-C * 03: 03: 01 and HLA-C * 14: 03 alleles were observed more frequently in the patient group(p = 0.007, p = 0.043, respectively)) and these alleles also were predisposed to a higher risk of disease. There was no statistically significant difference for HLA-DQB1 region. HLA DRB1 * 04: 05 allele (p = 0.013). was frequently observed in the patient group and predisposed to a higher risk of disease.

**Table 3.**
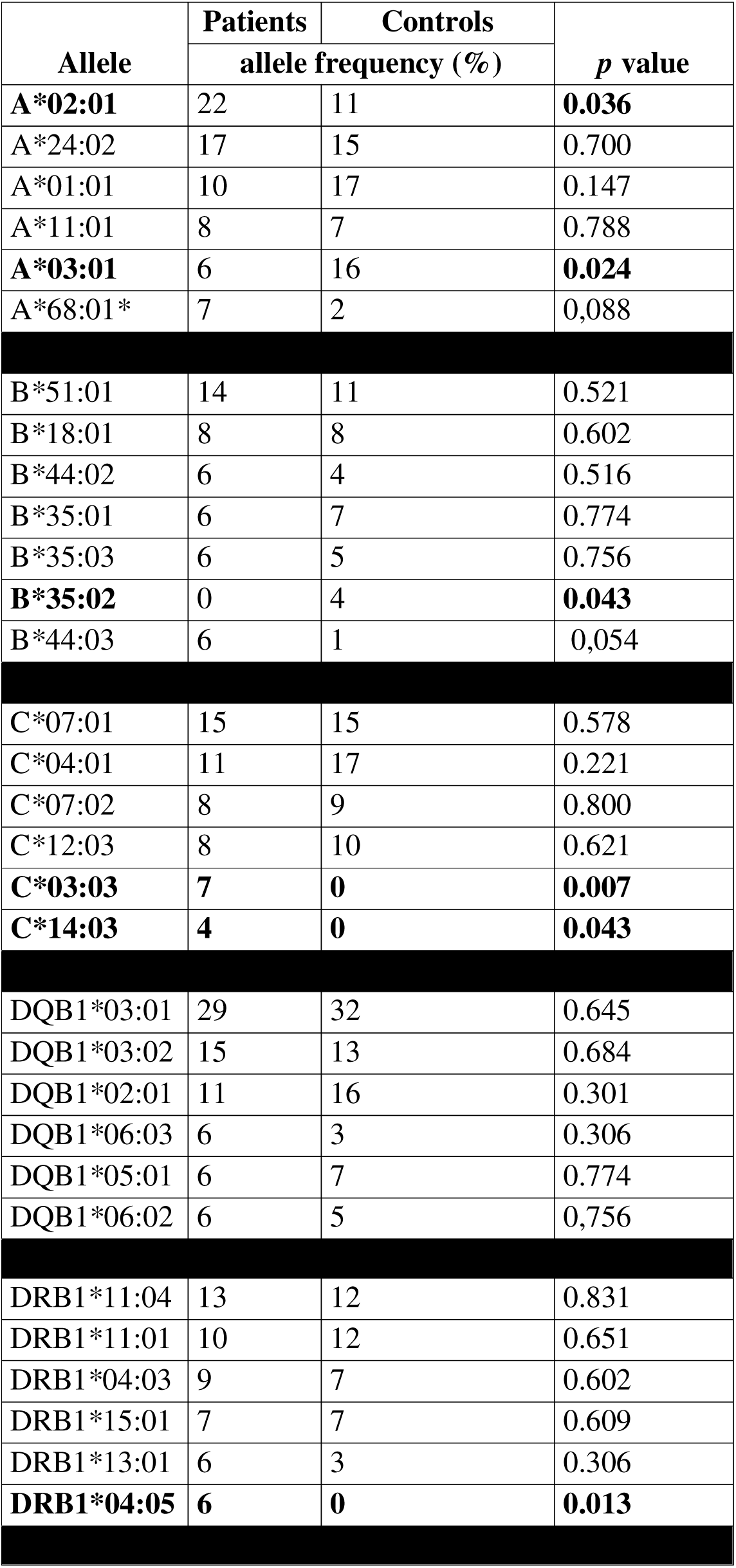
The six most frequent and OSA-related alleles of HLA-A, -B, -C, -DRB1, and - DQB1 loci.

| Allele | Patients | Controls | p value |
| --- | --- | --- | --- |
|  | allele frequency (%) |  |  |
| A*02:01 | 22 | 11 | 0.036 |
| A*24:02 | 17 | 15 | 0.700 |
| A*01:01 | 10 | 17 | 0.147 |
| A*11:01 | 8 | 7 | 0.788 |
| A*03:01 | 6 | 16 | 0.024 |
| A*68:01* | 7 | 2 | 0,088 |
| B*51:01 | 14 | 11 | 0.521 |
| B*18:01 | 8 | 8 | 0.602 |
| B*44:02 | 6 | 4 | 0.516 |
| B*35:01 | 6 | 7 | 0.774 |
| B*35:03 | 6 | 5 | 0.756 |
| B*35:02 | 0 | 4 | 0.043 |
| B*44:03 | 6 | 1 | 0,054 |
| C*07:01 | 15 | 15 | 0.578 |
| C*04:01 | 11 | 17 | 0.221 |
| C*07:02 | 8 | 9 | 0.800 |
| C*12:03 | 8 | 10 | 0.621 |
| C*03:03 | 7 | 0 | 0.007 |
| C*14:03 | 4 | 0 | 0.043 |
| DQB1*03:01 | 29 | 32 | 0.645 |
| DQB1*03:02 | 15 | 13 | 0.684 |
| DQB1*02:01 | 11 | 16 | 0.301 |
| DQB1*06:03 | 6 | 3 | 0.306 |
| DQB1*05:01 | 6 | 7 | 0.774 |
| DQB1*06:02 | 6 | 5 | 0,756 |
| DRB1*11:04 | 13 | 12 | 0.831 |
| DRB1*11:01 | 10 | 12 | 0.651 |
| DRB1*04:03 | 9 | 7 | 0.602 |
| DRB1*15:01 | 7 | 7 | 0.609 |
| DRB1*13:01 | 6 | 3 | 0.306 |
| DRB1*04:05 | 6 | 0 | 0.013 |

To summarize, HLA-A * 02: 01, HLA-C * 03: 03: 01, HLA-C * 14: 03, HLA DRB1 * 04: 05 alleles were found to be more frequently in OSA patients, but not in the controls (p=0.036, p=0.007, p=0.043 and 0.013, respectively). The allele frequencies of HLA-A * 03: 01 and HLA-B * 35: 02 were significantly higher in controls compared to OSA patients (p=0.024 and p=0.043).

## DISCUSSION

The search for a definitive genetic marker in Obstructive Sleep Apnea (OSA) has led many researchers to explore various candidates, yet the association with Human Leukocyte Antigen (HLA) genes remains notably understudied (Table 4).(5, 6) The pioneering investigation in this field was conducted by Yoshizawa et al. in Japan, who identified a higher frequency of the HLA-A2 allele among 32 male patients with OSA.(20) However, that study was unable to propose a definitive pathophysiological mechanism to explain this association. Furthermore, the significantly higher Body Mass Index (BMI) in their patient group introduced a confounding variable, making the direct HLA-OSA relationship somewhat controversial, as obesity itself is a primary risk factor for the disease.(20)

**Table 4.** Comparison of the results of our study and the studies in the literature evaluating the OSAS -HLA relationship.

|  | HLA genes |  |  |  |  | Patient (OSA)/control | Method | Relationship |
| --- | --- | --- | --- | --- | --- | --- | --- | --- |
|  | A | B | C | DR | DQ |  |  |  |
| Yoshizawa et al. (1993) | + | + | + | + |  | 32/32 | Standard lymphocyte micro-cytotoxicity test | HLA-A2 predisposing allele |
| Alzafer et al. (2003) | + | + | + | + | + | 45/28 | Standard lymphocyte micro-cytotoxicity test | HLA-A28, CW3 and DR15 predisposing alleles<br>HLA B27 and B13 protective allele |
| Brunetti et al. (2005) | + | + | + | + | + | 41OSA/32PH* | Standard lymphocyte micro-cytotoxicity test (Class I)<br>PCR/SSO (Class II) | HLA-B65 predisposing allele |
| Lee et al. (2005) | + | + |  | + |  | 25/200 | Standard lymphocyte micro-cytotoxicity test | HLA-A*11 and HLA-DRB1*09 predisposing alleles<br>DRB1*08 related with severity |
| Manzotte et al. (2013) |  |  |  |  | +<br>**DQB1*0602 | 894 | PCR-SSP | HLA-DQB1*0602-predisposing allele |
| Silva et al. (2015) |  |  |  | +<br>** DRB1 |  | 131/ 223 | PCR-SSP | DRB1*03-predisposing allele |
| Momany et al. (2017) |  |  |  | +<br>** DRB1*15 | +<br>** DQB1*0602 | 40/40 | PCR-SSP | HLA-DQB1*0602-predisposing allele HLA-DRB1*15 unrelated allele |
| Tepebaşı et al. (2018) |  |  |  | + |  | 64/32 | PCR-SSP | DRB1*03 predisposing allele<br>DRB1*08 related with severity |
| Present Study | + | + | + | + | + | 50/50 | NGS | HLA-A*02:01,<br>HLA-C*03:03:01,<br>HLA-C*14:03 and<br>HLA DRB1*04:05 predisposing alleles<br>HLA-A*03:01 and HLA-B*35:02, protective alleles |

\*PH: pulmonary hypertension, \*\*Only one allele evaluated in these studies.

Similarly, a study from Korea involving 25 patients (predominantly male) suggested associations with HLA-DRB1*09 and HLA-A11 alleles, while noting that HLA-DRB1*08 might be linked to disease severity.(21) While these early findings provided a starting point, they were often limited by small sample sizes, gender imbalance, and low-resolution typing methods. By utilizing high-resolution Next-Generation Sequencing (NGS) and a more balanced cohort, our study aims to clarify these discrepancies and provide a more robust genetic profile for OSA.

Beyond the Asian cohorts, research in other populations has yielded diverse results regarding the role of HLA Class II alleles in OSA. In a study conducted in Northern Portugal, the HLA-DRB1*03 allele was identified as a potential predisposing factor, particularly in obese male patients, further complicating the interplay between genetics and body mass.(22) The influence of HLA genes may extend beyond simple disease susceptibility to the modulation of sleep architecture itself. Manzetto et al. demonstrated that the HLA-DQB1*0602 allele was significantly associated with specific EEG patterns in patients with OSA, including differences in theta and low-delta wave activities during early sleep stages.(23) These findings suggest that this allele could serve as a biological marker affecting underlying sleep physiology. Supporting this, Momany et al. also confirmed a link between OSA and HLA-DQB1*0602, though they found no such association for the HLA-DRB1*15 allele.(24) In contrast, Brunetti et al., focusing on pediatric populations, observed a relationship between HLA-DRB1*15 and sleep-disordered breathing (SDB), while also noting a significantly higher expression of the HLA-B65 allele in these patients.(25) These disparate findings across various age groups and ethnicities underscore the necessity of high-resolution studies like ours to provide definitive clarity.

In the context of the Turkish population, only two studies to date have investigated the potential association between HLA genes and OSA. The first, published in 2003, reported a higher frequency of the HLA-DRB1*15 allele in the patient group.(26) More recently, a study in 2018 evaluated the frequency of HLA-DRB1 alleles and identified DRB1*03 as a predisposing factor, while noting that HLA-DRB1*08 was associated with disease severity.(27) However, these previous domestic studies were limited to specific Class II loci and utilized lower-resolution typing methods. By contrast, our study provides a more comprehensive genomic map by evaluating HLA-A, -B, -C, -DRB1, and - DQB1 loci simultaneously using high-resolution Next-Generation Sequencing (NGS). The comparative results of international research, previous domestic studies, and our current findings are summarized in Table 4. This comprehensive comparison highlights that while some alleles like DRB1*03 show consistency in certain populations, the high-resolution data provided by our NGS analysis reveals novel associations, such as HLA-A*02:01 and HLA-DRB1*04:05, which were not previously highlighted in the Turkish OSA literature.

Among the existing literature, a comprehensive HLA region analysis comparable in detail to our study was only performed by Yoshizawa et al., Brunetti et al., and Alzafer et al. However, significant methodological differences exist between these studies and ours. Yoshizawa and Alzafer utilized the standard lymphocyte microcytotoxicity test, a serological method that identifies antigens at a lower resolution.(20, 26) Brunetti et al. employed a hybrid approach, using the lymphocyte microcytotoxicity test for Class I antigens and the PCR/SSO (Sequence-Specific Oligonucleotide) method for Class II antigens.(25) By contrast, the present study utilized Next-Generation Sequencing (NGS), which represents the current gold standard for HLA typing. Unlike serological tests or earlier PCR-based methods (SSO/SSP) that often yield ambiguous results or low-resolution data, NGS allows for unambiguous, high-resolution allele identification at the four-digit level. This technological precision is likely the reason we were able to identify specific associations, such as HLA-A*02:01 and HLA-DRB1*04:05, which may have been missed or characterized only broadly in earlier investigations.

Serological methods are inherently limited, as they can identify only a very small fraction of the vast spectrum of HLA alleles. Similarly, though more advanced, SSO or SSP methods are restricted to detecting previously known alleles and are generally suitable only for two-digit resolution of HLA polymorphic loci.(10, 11) In the present study, the application of Next-Generation Sequencing (NGS) methodology facilitated a high-resolution (4 to 6-digit) evaluation of these genes, enabling the identification of all probable alleles with unprecedented precision. It is also critical to consider the temporal context of HLA research. The pioneering work of Yoshizawa et al. was conducted approximately 33 years ago, and Brunetti et al.’s research took place over a decade ago.(20, 25) While their findings remain foundational, they were limited by older techniques and a significantly smaller database of identified HLA polymorphisms compared to the robust, cumulative databases available today. Our high-resolution analysis revealed a statistically significant difference in the frequencies of the HLA-A*02:01 and HLA-A*03:01 alleles (p = 0.036 and p = 0.024, respectively) between the OSA and control groups. Specifically, HLA-A*02:01 was more prevalent in the patient group, suggesting its role as a predisposing factor for the disease. Conversely, HLA-A*03:01 was found more frequently in the control group, indicating a potential protective effect against the development of OSA.

In the historical development of OSA genetics, our findings both validate and significantly refine earlier observations. While Yoshizawa et al. previously reported an association between the HLA-A*02 allele and OSA, their serological approach lacked the resolution to distinguish between specific subtypes.(20) By contrast, the high-resolution four-digit NGS assessment employed in our study enabled us to pinpoint HLA-A*02:01 as the precise subgroup predisposing to the disease. Most importantly, our granular analysis revealed a divergent role for other alleles within the same locus; while HLA-A*02:01 increased risk, HLA-A*03:01 was identified as a protective factor. This level of detail explains why studies using lower-resolution methods, such as that by Brunetti et al., may have failed to identify a significant relationship within the HLA-A region.(25) Furthermore, our findings differ from the Korean cohort studied by Lee et al., who identified HLA-A*11 as a predisposing factor, highlighting how high-resolution typing can uncover population-specific genetic variations that remain hidden in broader antijen-level investigations.(21)

Our analysis of the HLA-B region revealed that the HLA-B*35:02 allele appears to exert a protective effect against OSA, as it was observed with significantly greater frequency in the control group compared to patients. This finding is particularly noteworthy when compared to the existing literature. For instance, Brunetti et al. identified an association between the HLA-B*65 allele and OSA.(25) However, such discrepancies are expected given that the HLA locus is a highly polymorphic gene region characterized by profound ethnic differences. In fact, the protective role of HLA-B*35:02 identified in our cohort may be unique to the Turkish population, as previous domestic studies in Turkey did not report such an association.(26, 27) Furthermore, our results for the HLA-B*44:03 allele yielded a p-value of 0.054. While this was technically considered statistically insignificant, it represents a notable borderline value. This suggests that the HLA-B*44:03 allele may indeed play a role in OSA susceptibility, a possibility that warrants further investigation in larger cohorts to achieve greater statistical power.

One of the most striking findings of our study relates to the HLA-C region, where we identified a statistically significant association for the HLA-C*03:03:01 and HLA-C*14:03 alleles (p = 0.007 and p = 0.043, respectively). Both alleles were observed with a significantly higher frequency in the OSA patient group, suggesting that they may serve as potent predisposing factors for the disease. To the best of our knowledge, no such association between these specific HLA-C alleles and OSA has been previously reported in the literature. Most prior studies have focused predominantly on the HLA-A and HLA-DRB1 regions, often overlooking the potential contribution of the HLA-C locus. Our discovery of these novel markers highlights the advantage of using high-resolution NGS to screen the entire HLA complex, and it suggests that the HLA-C region may play a more central role in the genetic susceptibility to OSA than previously recognized.

Regarding the HLA Class II region, our analysis of the HLA-DQB1 locus did not reveal any statistically significant associations. This stands in contrast to the findings of Manzotte et al. and Momany et al., who previously reported that the HLA-DQB1*06:02 allele was a significant predisposing factor for OSA.(23, 24) The absence of this association in our cohort further emphasizes the role of ethnic heterogeneity in the genetic architecture of OSA, suggesting that the DQB1*06:02 marker may not be a universal predictor across all populations. However, a significant association was identified within the HLA-DRB1 region. Specifically, the HLA-DRB1*04:05 allele was observed with a significantly higher frequency in the patient group compared to controls (p = 0.013), indicating its role as a novel predisposing factor for OSA. While previous domestic and international studies, such as those by Tepebaşı et al. and Silva et al., have highlighted the DRB1*03 and DRB1*09 alleles as susceptibility markers,(22, 27) there is no prior report in the literature linking the DRB1*04:05 allele to OSA. This discovery of a novel genetic marker, made possible through high-resolution NGS, provides a new target for future research into the immunogenetic mechanisms underlying the disease.

### Limitations

The primary limitation of the present study is the relatively small sample size in both the patient and control groups. High-resolution NGS-based HLA typing remains a high-cost methodology, and the funding resources of our center necessitated a more focused cohort. Despite this limitation, our study yielded significant and novel findings that contribute to the current understanding of the genetic basis of OSA. However, we acknowledge that these results should be interpreted as foundational, and larger, multi-center studies with greater statistical power are essential to further elucidate the complex relationship between HLA genes and OSA.

### CONCLUSIONS AND RECOMMENDATIONS

In the present study, a comprehensive analysis of the HLA-A, -B, -C, -DQB1, and -DRB1 loci using Next-Generation Sequencing (NGS) revealed significant associations between specific HLA alleles and Obstructive Sleep Apnea (OSA) pathogenesis. Our findings identify HLA-A*02:01, HLA-C*03:03:01, HLA-C*14:03, and HLA-DRB1*04:05 as novel disease-predisposing alleles. Conversely, HLA-A*03:01 and HLA-B*35:02 were observed at significantly higher frequencies in healthy controls, suggesting a potential protective role against the development of OSA.

Given the highly polymorphic nature and profound ethnic diversity of the Major Histocompatibility Complex (MHC) region, these previously unreported associations provide critical insights that may be specific to our geographical region. While previous studies have hinted at an HLA-OSA link, the overall number of investigations—both globally and within our country—remains remarkably low. Our findings underscore the urgent need for large-scale, multicenter studies to further elucidate this genetic concordance.

Furthermore, it is important to emphasize that this study utilized the most sensitive DNA sequencing technology currently available. To the best of our knowledge, this is the first study to investigate the HLA-OSA relationship using the NGS methodology. Although NGS is more resource-intensive than traditional methods, its ability to provide four-to-six-digit resolution ensures an unparalleled level of precision in allele detection. Considering the well-known ethnic variations in the HLA region, widespread adoption of NGS in different global populations will be essential to fully mapping the immunogenetic landscape of OSA.

### Declaration of Generative AI in Scientific Writing

During the preparation of this work, the authors used Gemini 3 Flash (Google, Mountain View, CA, USA) for linguistic assistance and academic phrasing. Following the use of this tool, the authors independently reviewed, refined, and edited the manuscript to ensure scientific accuracy. The authors take full responsibility for the final content and integrity of the publication.

## Data Availability

The authors confirm that the data supporting the findings of this study are available within the article [and/or] its supplementary materials.

## Acknowledgements

The authors would like to thank the clinical and technical staff of the Departments of Chest Diseases and Medical Genetics at Necmettin Erbakan University for their invaluable administrative and technical support. This study was supported by the Scientific Research Projects Coordination Unit (BAP) of Necmettin Erbakan University (Project no: 171218006).

## Author Contributions

**Conceptualization:** Zamani AG, Zamani A.

**Data curation:** Zamani AG, Yosunkaya S, Korkmaz C.

**Formal analysis:** Zamani AG, Arslan AB, Korkmaz C.

**Funding acquisition:** Zamani AG.

**Investigation:** Zamani AG, Yosunkaya S, Vatansev H.

**Methodology:** Zamani AG, Yosunkaya S, Vatansev H, Korkmaz C, Yildirim MS.

**Project administration:** Zamani AG, Zamani A.

**Resources:** Zamani AG, Yosunkaya S, Vatansev H.

**Software:** Zamani AG, Korkmaz C, Arslan AB.

**Supervision:** Zamani AG.

**Validation:** Zamani AG, Yosunkaya S, Vatansev H, Korkmaz C.

**Visualization:** Zamani AG, Korkmaz C, Zamani A, Yildirim MS.

**Writing - original draft:** Zamani AG, Arslan AB, Korkmaz C, Zamani A.

**Writing - review & editing:** Zamani AG, Yildirim MS, Korkmaz C, Zamani A.

